# Stable IgG-antibody levels in patients with mild SARS-CoV-2 infection

**DOI:** 10.1101/2021.06.16.21258960

**Authors:** Thomas Åkerlund, Katherina Zakikhany, Charlotta Löfström, Evelina Lindmark, Henrik Källberg, Ulla Elofsson, Karin Cederbrant, Erik Nygren, Anders Kallin, Nina Lagerqvist, Peter Nilsson, Sophia Hober, Anna Ridderstad Wollberg, Åsa Szekely Björndal

**Author notes:** Corresponding author: Thomas Åkerlund, Department of Microbiology, Public Health Agency of Sweden, Solna Sweden.

## Abstract

More knowledge regarding persistence of antibody response to SARS-CoV-2 infections in the general population with mild symptoms is needed. We measured and compared levels of SARS-CoV-2 spike- and nucleocapsid-specific IgG-antibodies in serum samples from 145 laboratory-confirmed COVID-19 cases and 324 non-cases. The IgG-antibody levels against the spike protein in cases were stable over the time-period studied (14 to 256 days), while antibody levels against the nucleocapsid protein decreased over time.

## Introduction

The ongoing pandemic of the severe acute respiratory syndrome coronavirus 2 (SARS-CoV-2) and its associated respiratory disease, coronavirus disease 2019 (COVID-19), poses a major and unprecedented threat to global public health (1).

To date, serological tests are widely used to screen for antibodies made in response to a SARS-CoV-2 infection. The antibody responses to SARS-CoV-2 infection are directed against multiple viral antigens including the spike and the nucleocapsid protein (2). Antibodies against the spike-S1 protein, more specifically, the receptor-binding domain, have shown virus-neutralizing activity (3).

Understanding the antibody kinetics over prolonged periods is of crucial importance especially with the ongoing roll out of general vaccination and immunity passports being discussed. Recent reports describe detectable IgG levels in approximately 80-90% of infected COVID-19 patients 4-8 months post infection (4-8). Several studies have included health-care workers with confirmed COVID-19 (9, 10), whose immune response may differ from that of the general population. In this context, there is a need for better understanding of antibody responses in the general population with largely mild symptoms.

Here, we characterize 469 serum samples collected from the general population in Sweden during October to December 2020. The samples were collected to establish a well-characterized national serum panel consisting of SARS-CoV-2 antibody-positive and negative samples for performance evaluation and quality assurance of serological tests. We assessed the antibody levels against SARS-CoV-2 nucleocapsid and spike proteins in sera from 145 COVID-19 patients 14 to 256 days after diagnosis.

## Materials and methods

### Recruitment of study participants

Study participants were recruited from three Swedish Counties via internet-based advertisement. The advertisement was specifically directed towards persons who either had or had not contracted COVID-19 during 2020. A web-based questionnaire was used to collect information on basic demographics and anamnesis concerning COVID-19 (Figure S1). The purpose of the questionnaire was to pre-screen for candidates likely to donate either antibody-positive or negative serum samples, as well as to allow for weighing of samples regarding gender, age, county of residence (county 1-3) and symptoms of respiratory tract infection (RTI) other than COVID-19. Of 5444 persons completing the questionnaires, 780 individuals were selected for serum sampling according to the criteria mentioned above. Of these, 484 donated serum for the study (Table S1). Written consent was obtained from all study-participants. At the time of serum sampling, vaccination campaigns had not yet started in Sweden.

### Serum sampling

Venous blood (5-40 mL) was collected into Serum Sampling Tubes (SST BD Vacutainer) on county level and transported at ambient temperature to regional laboratories for centrifugation and isolation of serum. Serum sample aliquots were subsequently shipped to RISE Research Institutes of Sweden for further analysis.

### Case definitions

Patient-cases were defined as study participants with a laboratory-confirmed SARS-CoV-2 infection, i.e. with positive real-time reverse transcription PCR test (rRT-PCR), as reported in the national Swedish SmiNet database (electronic monitoring system for notifications received in accordance with the Communicable Diseases Act). For data analysis, dates for rRT-PCR-diagnostics were primarily used, and if missing, disease notification dates were used.

Non-cases were defined as study participants with no reported laboratory-confirmed SARS-CoV-2 infection (i.e. rRT-PCR test) as per recording in the SmiNet database up to the serum sampling date.

Of 484 participants, 15 were excluded due to incomplete personal identification number, missing serum samples, or if serum sampling had occurred within 14 days of disease onset.

One follow-up search in the SmiNet database for possible reinfections and/or new infections for the cohort was performed on April 12, 2021.

### Antibody testing

Sera were tested for the presence of IgG antibodies against SARS-CoV-2 at the Science for Life Laboratory (SciLifeLab) in Solna, Sweden. Testing was performed using a semi-quantitative multiplex bead-based serologic assay for detection of antibodies to two different SARS-CoV-2 viral proteins, the C-terminal part of the nucleocapsid protein (NCP-C) and the soluble spike glycoprotein (spike-foldon) (11). The sensitivity and specificity of the method was specified as 99.7% (95% CI: 98.3-100%) and 100% (95% CI: 99.8-100%), respectively. Results were obtained as fluorescence intensity in a FlexMap3D instrument (Luminex) and are here given as arbitrary units (AU). Each run included twelve negative and four positive reference samples. Cut-off was determined as the “mean + 6 × SD” of the twelve negative reference samples. Seropositivity was defined as AU above the cut-off for both the nucleocapsid and spike protein. The median AU value of the four positive samples was used for normalization between assays. Three separate runs were performed, and the values were normalized against the run with the largest number of samples. Testing was performed blinded, i.e. without disclosing any case information of the samples.

### Statistical analysis

To compare groups, i.e. cases vs. non-cases and cases only regarding severity (hospital vs. ill at home) we performed pairwise t-test, Mann-Whitney U-test and ANCOVA. ANCOVA was used to allow for correction of variability of gender, age and time between diagnosis (positive rRT-PCR) and serum sampling. We only present P-values based on ANCOVA for group comparisons. Furthermore, we estimated Pearsons correlation coefficient for the log of nucleocapsid and spike antibody levels (AU) in cases. R version 3.6.2 was used for all analysis.

### Ethical approval and financing

Ethical approval for this study was obtained from the Swedish Ethical Review Board: Dnr 2020-03584. This project was exclusively funded by the Public Health Agency of Sweden via an assignment (S2020_05026) from the Ministry of Health and Social Affairs, Government of Sweden.

## Results

### Demographics and clinical characteristics

Serum samples from the 469 included study participants were collected between October 30 and December 8, 2020. Of all participants, 145 were found to be laboratory-confirmed COVID-19-patients in the national registry system (SmiNet) and subsequently classified as patient-cases. All patient-cases had sampling/diagnosis dates between March 6 and November 9, 2020 and had until April 21, 2021 only one record in SmiNet. Thus, there were no patient-cases with a recorded re-infection. The remaining 324 participants had no record in SmiNet and were subsequently denoted as non-cases (Table 1). Of note, 28/324 non-cases (8.6%) were registered with a COVID-19 diagnosis in the follow-up search on April 12, 2021.

**Table 1.** Age, gender and self-reported symptoms of patient-cases and non-cases.

|  | Patient-cases (%) | Non-cases (%) | Total (%) |
| --- | --- | --- | --- |
| **N** | 145 (31) | 324 (69) | 469 (100) |
| **Gender** |  |  |  |
| Female | 89 (61) | 181 (56) | 270 (58) |
| Male | 56 (39) | 143 (44) | 199 (42) |
| **Age group** |  |  |  |
| 18-29 | 14 (10) | 45 (14) | 59 (13) |
| 30-44 | 39 (27) | 78 (24) | 117 (25) |
| 45-59 | 70 (48) | 114 (35) | 184 (39) |
| ≥60 | 22 (15) | 87 (27) | 109 (23) |
| **Symptoms of respiratory-tract infections during 2020\*** |  |  |  |
| Yes | 115 (79) | 127 (39) | 242 (52) |
| No† | 30 (21) | 197 (61) | 227 (48) |
| **Severity of symptoms‡** |  |  |  |
| Mild | 123 (85) | 52 (16) | 175 (37) |
| Moderate | 16 (11) | 3 (0.9) | 19 (4.1) |
| Severe | 5 (3.4) | 0 (0) | 5 (1.1) |
| No data | 1 (0.7) | 269 (83) | 270 (58) |

\*Symptoms of respiratory tract infections and severity of disease were extracted from the questionnaires.
†Of those that reported no RTI-symptoms, 29/30 of patient-cases and 12/197 of non-cases, respectively, reported symptoms that they suspected were due to COVID-19.
‡The question of severity of symptoms applied only to those who answered “Yes” on the question “Did you have symptoms during 2020 that you know, or suspect were due to COVID-19” (n = 144 for cases, and n = 55 for non-cases). Mild disease was defined as staying at home while being ill, moderate as hospital admission and severe as ICU admission.

Most patient-cases, 85%, self-reported severity of illness as mild (Table 1) and symptoms were dominated by influenza-like symptoms (Table S2). About 39% of non-cases reported symptoms of respiratory-tract infection during 2020 and furthermore, 17% of non-cases reported that they had or suspected to have had contracted COVID-19 during 2020 (Table 1).

### IgG antibody levels in patient-cases and non-cases

About 95% of patient-cases (137/145) were classified as seropositive, i.e. serum samples showed IgG antibody levels above the threshold for both the nucleocapsid and spike proteins (Table 2). Of the eight seronegative samples, four had antibodies against either spike or nucleocapsid and another four had no detectable antibodies against any of the included antigens. The four non-reactive samples showed background levels in the same range as the majority of non-case samples for the spike protein (Figure 1) and the fluorescence intensity levels showed a discontinuous distribution (Figure 2). These four non-reactive samples were thus considered as outliers and were excluded for the analyses. There were no significant differences in seropositivity or antibody levels between age groups (Figure S2) or gender (not shown).

**Table 2.** IgG-antibodies detected in serum samples of patient-cases and non-cases.

| Reactive IgG antibody<br>against | Patient-cases<br>(n=145) | % (95% CI) | Non-cases<br>(n=324) | % (95% CI) |
| --- | --- | --- | --- | --- |
| Spike | 140 | 96.6 (92.2-98.5) | 21 | 6.5 (4.3-9.7) |
| Nucleocapsid | 138 | 95.2 (90.4-97.6) | 18 | 5.6 (3.6-8.6) |
| Spike and Nucleocapsid | 137 | 94.5 (89.5-97.2) | 14 | 4.3 (2.6-7.1) |
| None | 4 | 2.8 (1.1-6.9) | 299 | 92.3 (88.9-94.7) |

**Figure 1.**
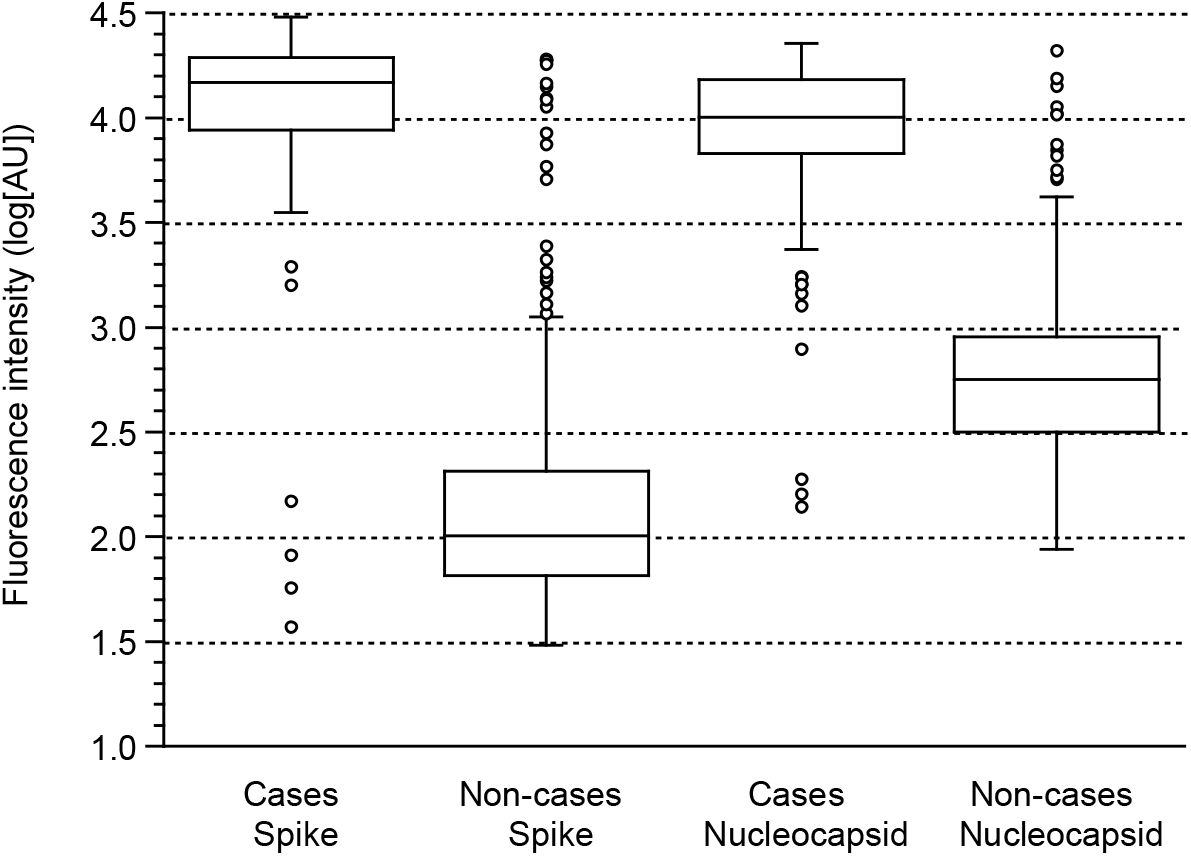
Antibody levels against viral spike and nucleocapsid proteins in patient-cases (n = 145) and non-cases (n = 324), respectively. Boxes include values between the first (Q1) and third (Q3) quartile, whiskers limits shows values within Q1-IQR×1.5 and Q3+IQR×1.5. Outliers are defined as values outside the whisker limits and median values are indicated by a line. IQR: Interquartile range.

**Figure 2.**
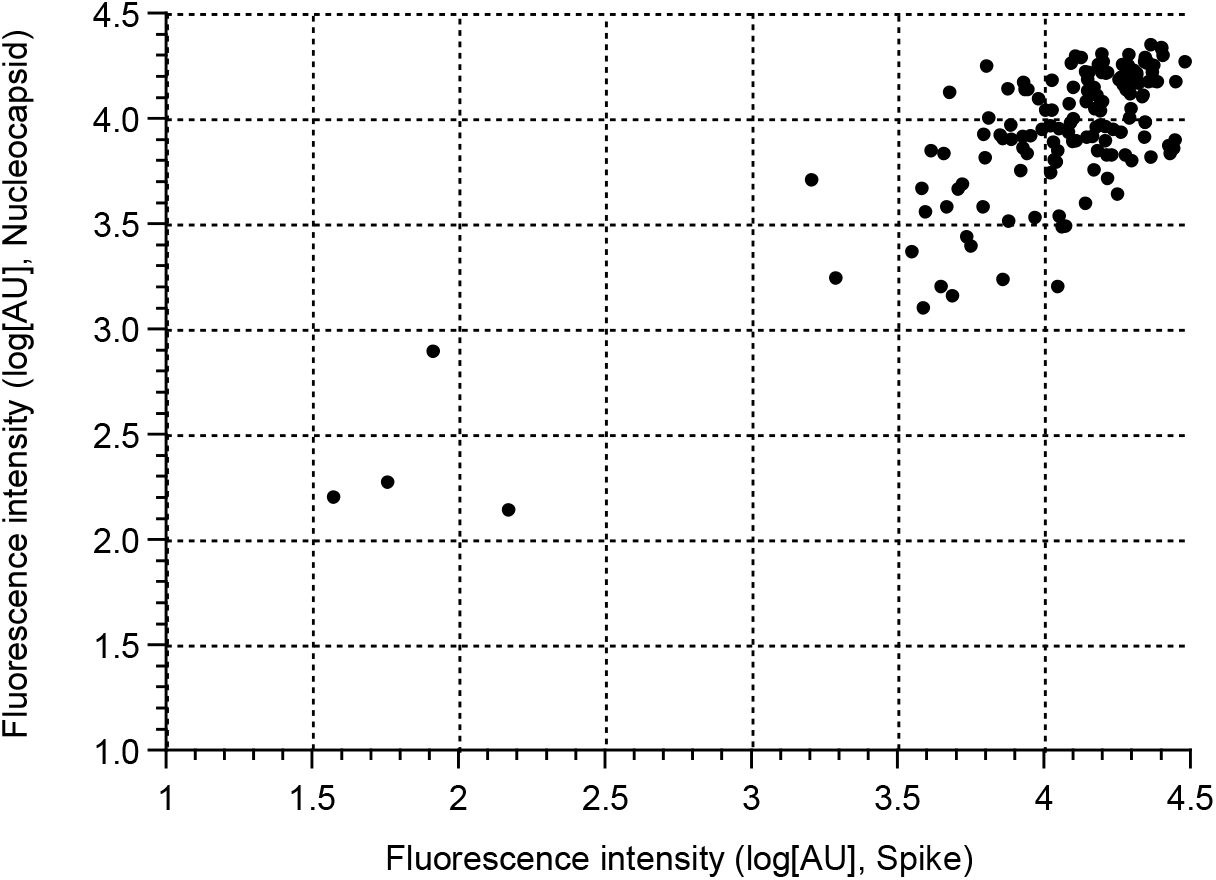
Correlation of IgG antibody levels against nucleocapsid and spike proteins among patient-cases (n=145).

Fourteen non-cases (4%) were classified as seropositive (Table 2). Among the 55 non-cases that self-reported to have contracted COVID-19 during 2020, 10 tested seropositive (18%) and 45 tested negative (82%). Geographically, County 1 had a higher proportion of seropositive samples among non-cases than the other two counties (Table S1).

### IgG antibody levels and severity of disease

Among patient-cases, antibody levels against the spike protein were higher in patients with moderate to severe disease compared to patients with mild disease (P = 0.004, ANCOVA, Figure 3). A similar association was found for antibody levels against the nucleocapsid protein (P = 0.007, ANCOVA).

**Figure 3.**
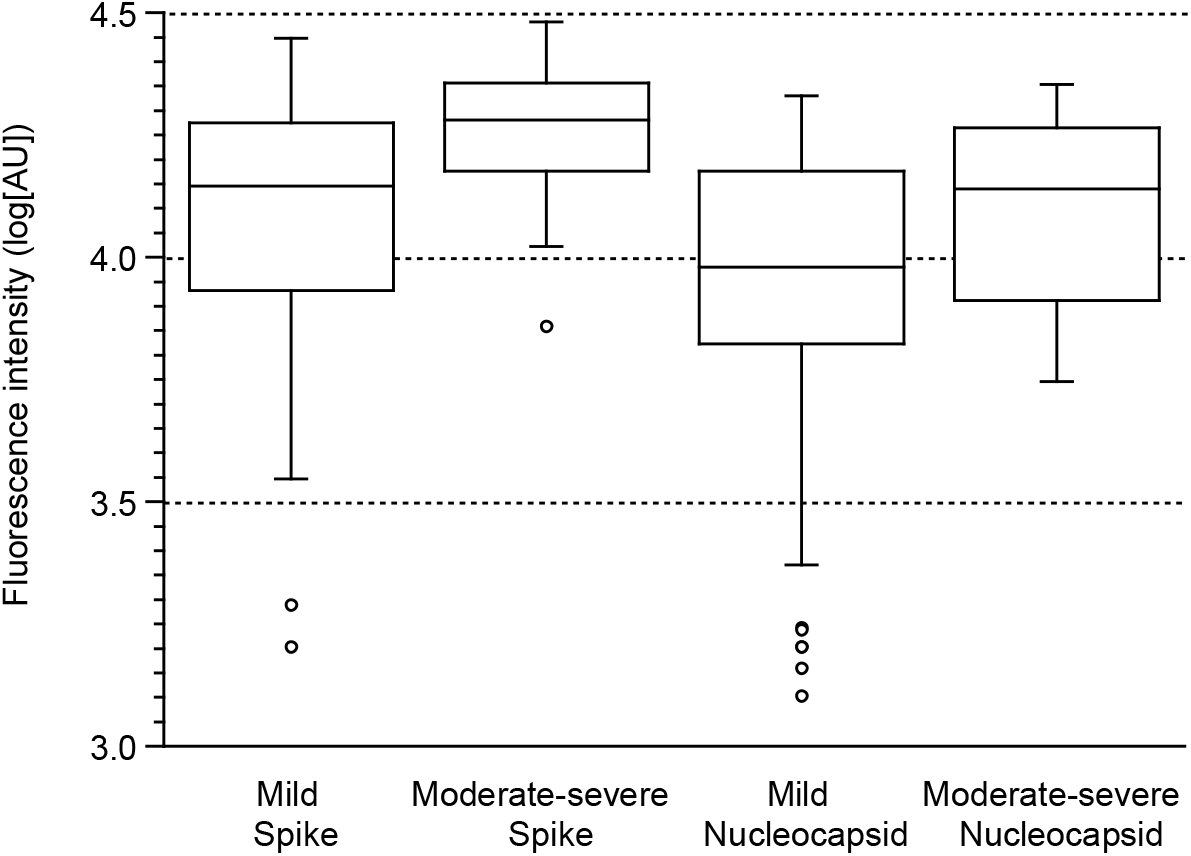
Antibody levels in patient-cases with mild vs. moderate-severe disease (n = 120 vs. n = 21, respectively, four outliers removed). For explanation of boxes, see legend to Figure 1.

### IgG antibody levels in patient-cases in relation to time after diagnosis

For 141 patient-cases (excluding the four outliers), disease notification dates were recorded between March 6 and November 9, 2020, with a majority (30%) recorded in June, followed by 17% in May and October, respectively. The median time between disease notification date and serum sampling was 156 days (range 15-256 days). Four samples were collected 15–20 days after disease notification and 137 samples between 21–256 days. For the 4/141 samples that were classified as seronegative, three samples tested positive for antibodies against the spike protein and one sample tested positive for antibodies against the nucleocapsid protein. These samples were collected at day 160, 176, 207, and 232 after diagnosis.

There were no significant differences in IgG antibody levels against the spike protein over time (Figure 4, R = -0.059, P = 0.49). Conversely, antibody levels against the nucleocapsid protein were lower for patients diagnosed earlier during spring compared to those in the autumn (R = -0.19; P = 0.02).

**Figure 4.**
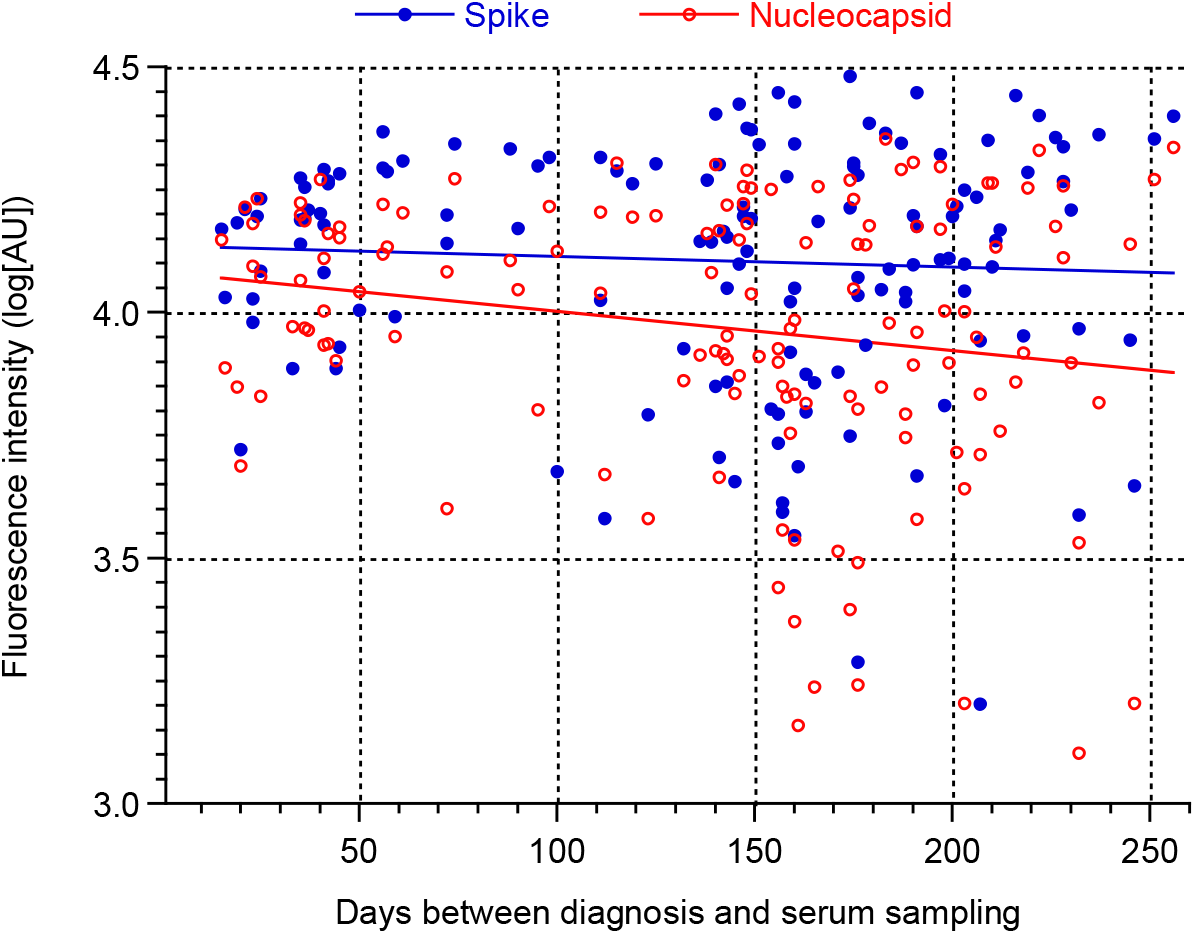
Correlation of antibody levels among patient-cases and time between disease notification and serum sampling (n = 141, 4 outliers removed).

## Discussion

We describe the IgG antibody levels in 145 serum samples from patients with a confirmed COVID-19 diagnosis between March and November 2020 in Sweden. The 145 patient samples were part of 469 serum samples, collected with the purpose to establish a national serum panel for validation and quality assessment of serological assays.

Our data showed that 95% of patient-cases seroconverted after a laboratory-confirmed infection with SARS-CoV-2, and there was no difference with respect to age group or gender. Early studies reported 15-90% seroconversion rate in COVID-19 patients, with lower rates among asymptomatic patients (12, 13). One explanation for the varying results are performance differences among serological methods (14-16). Despite the high performance of the method used in this study, eight samples were classified as seronegative. Whereas four patient-cases had detectable IgG levels against one of the two antigens, the remaining four were distinguished by having background fluorescence levels similar to those of the negative controls. Although not conclusive, these outliers most likely represent false positive rRT-PCR diagnostic results. Large scale diagnostic testing in low prevalence settings increases the risk for false positives (17), and highlights the need for careful interpretation of serological non-responders as well as adherence to the case definition of putative reinfections (18).

The levels of IgG antibodies against the spike and nucleocapsid proteins were higher in patients with moderate to severe disease than in patients with mild disease. Reports have shown various results regarding the antibody levels in patients with severe and mild disease (16, 19, 20), which may be due to timing of antibody response in relation to the phase of the infection (21).

One strength of this study is that samples were collected from the general population, covering patients with mostly mild disease. The results indicate that the antibody levels, particularly against the spike protein, remain at detectable levels for more than eight months. This is in agreement with other studies showing a sustained antibody response, e.g. 6-9 months after infection (4, 5, 7, 22-24). Neutralizing antibodies are evidently important for protection against disease (25), and protection over longer time-periods depends on the evolution of memory B cells (26). In this context, it is interesting to note that only one episode of COVID-19 per case was registered between March 6, 2020 and April 12, 2021, while 9% of the non-cases were diagnosed with COVID-19 during the 3.5 months that passed after serum sampling.

Due to the unknown number of undiagnosed SARS-CoV-2 infections in the population, one challenge of our approach was to collect true negative serum samples. Twelve of the 55 non-cases who self-reported a suspected COVID-19 infection also reported positive rRT-PCR-test results in the questionnaires, despite not being registered in the national registry system. Possible explanations for this discrepancy are that they had received their diagnoses abroad, or that there was a misinterpretation of the questionnaire. Nevertheless, excluding those who had reported being tested positive for SARS-CoV-2 with rRT-PCR among the non-cases, we found 7/312 seropositive samples among the non-cases. Six of these were from County 1 which had the highest surge of COVID-19 during the first months of the pandemic in Sweden. About 9% of non-cases in County 1 had detectable antibody levels against either the spike or nucleocapsid protein, compared to 2% in the other two Counties (Table S3). Due to the non-random selection and bias in the sampling algorithm used here, the result is most likely an underestimate of the proportion of undiagnosed cases in the population.

The well-characterized panel of serum samples presented here will be valuable for diagnostic performance and quality assessments of current and new serological assays.

## Data Availability

All data are included in the manuscript and appendix. Data is not available at other URLs.

## Acknowledgments

We thank all voluntary participants for donating blood. We also want to thank the health care centres and regional laboratories for blood sampling and serum preparation as well as the SciLifeLab Autoimmunity and Serology profiling facility for analysis of serum samples. Jenny Belin, Carolina Sandman, Jenny Lindahl and Ingela Persson, RISE Research Institutes of Sweden, are acknowledged for setting up the quality management system handling the serum samples, technical assistance and for providing valuable advice on sampling. We are grateful to Karin Tegmark-Wisell, Charlotta Nilsson and Jonas Klingström, Public Health Agency of Sweden, and Magnus Larsson, RISE Research Institute of Sweden, for reading and giving valuable input to the manuscript.

## Notes

### Competing Interest Statement

The authors have declared no competing interest.

### Author Declarations

Ethical approval for this study was obtained from the Swedish Ethical Review Board: Dnr 2020-03584.

## References

1. Cucinotta D, Vanelli M. WHO Declares COVID-19 a Pandemic. Acta Biomed. 2020 Mar 19;91(1):157–60.

2. Li D, Li J. Immunologic Testing for SARS-CoV-2 Infection from the Antigen Perspective. J Clin Microbiol. 2021 Apr 20;59(5).

3. Liu L, Wang P, Nair MS, Yu J, Rapp M, Wang Q, et al. Potent neutralizing antibodies against multiple epitopes on SARS-CoV-2 spike. Nature. 2020 Aug;584(7821):450–6.

4. Sherina N, Piralla A, Du L, Wan H, Kumagai-Braesch M, Andrell J, et al. Persistence of SARS-CoV-2-specific B and T cell responses in convalescent COVID-19 patients 6-8 months after the infection. Med (N Y). 2021 Mar 12;2(3):281–95 e4.

5. Anand SP, Prevost J, Nayrac M, Beaudoin-Bussieres G, Benlarbi M, Gasser R, et al. Longitudinal analysis of humoral immunity against SARS-CoV-2 Spike in convalescent individuals up to 8 months post-symptom onset. bioRxiv. 2021 Jan 25.

6. Abayasingam A, Balachandran H, Agapiou D, Hammoud M, Rodrigo C, Keoshkerian E, et al. Long-term persistence of RBD(+) memory B cells encoding neutralizing antibodies in SARS-CoV-2 infection. Cell Rep Med. 2021 Apr 20;2(4):100228.

7. den Hartog G, Vos ERA, van den Hoogen LL, van Boven M, Schepp RM, Smits G, et al. Persistence of antibodies to SARS-CoV-2 in relation to symptoms in a nationwide prospective study. Clin Infect Dis. 2021 Feb 24.

8. Gudbjartsson DF, Norddahl GL, Melsted P, Gunnarsdottir K, Holm H, Eythorsson E, et al. Humoral Immune Response to SARS-CoV-2 in Iceland. N Engl J Med. 2020 Oct 29;383(18):1724–34.

9. Lumley SF, Wei J, O’Donnell D, Stoesser NE, Matthews PC, Howarth A, et al. The duration, dynamics and determinants of SARS-CoV-2 antibody responses in individual healthcare workers. Clin Infect Dis. 2021 Jan 6.

10. Dan JM, Mateus J, Kato Y, Hastie KM, Yu ED, Faliti CE, et al. Immunological memory to SARS-CoV-2 assessed for up to 8 months after infection. Science. 2021 Feb 5;371(6529).

11. Rudberg AS, Havervall S, Manberg A, Jernbom Falk A, Aguilera K, Ng H, et al. SARS-CoV-2 exposure, symptoms and seroprevalence in healthcare workers in Sweden. Nat Commun. 2020 Oct 8;11(1):5064.

12. Long QX, Tang XJ, Shi QL, Li Q, Deng HJ, Yuan J, et al. Clinical and immunological assessment of asymptomatic SARS-CoV-2 infections. Nat Med. 2020 Aug;26(8):1200–4.

13. Wellinghausen N, Plonne D, Voss M, Ivanova R, Frodl R, Deininger S. SARS-CoV-2-IgG response is different in COVID-19 outpatients and asymptomatic contact persons. J Clin Virol. 2020 Sep;130:104542.

14. Harris RJ, Whitaker HJ, Andrews NJ, Aiano F, Amin-Chowdhury Z, Flood J, et al. Serological surveillance of SARS-CoV-2: Six-month trends and antibody response in a cohort of public health workers. J Infect. 2021 Mar 22.

15. Marklund E, Leach S, Axelsson H, Nystrom K, Norder H, Bemark M, et al. Serum-IgG responses to SARS-CoV-2 after mild and severe COVID-19 infection and analysis of IgG non-responders. PLoS One. 2020;15(10):e0241104.

16. Wolf J, Kaiser T, Pehnke S, Nickel O, Lubbert C, Kalbitz S, et al. Differences of SARS-CoV-2 serological test performance between hospitalized and outpatient COVID-19 cases. Clin Chim Acta. 2020 Dec;511:352–9.

17. Surkova E, Nikolayevskyy V, Drobniewski F. False-positive COVID-19 results: hidden problems and costs. Lancet Respir Med. 2020 Dec;8(12):1167–8.

18. Yahav D, Yelin D, Eckerle I, Eberhardt CS, Wang J, Cao B, et al. Definitions for coronavirus disease 2019 reinfection, relapse and PCR re-positivity. Clin Microbiol Infect. 2021 Mar;27(3):315–8.

19. Wu F, Liu M, Wang A, Lu L, Wang Q, Gu C, et al. Evaluating the Association of Clinical Characteristics With Neutralizing Antibody Levels in Patients Who Have Recovered From Mild COVID-19 in Shanghai, China. JAMA Intern Med. 2020 Oct 1;180(10):1356–62.

20. Sun B, Feng Y, Mo X, Zheng P, Wang Q, Li P, et al. Kinetics of SARS-CoV-2 specific IgM and IgG responses in COVID-19 patients. Emerg Microbes Infect. 2020 Dec;9(1):940–8.

21. Li K, Huang B, Wu M, Zhong A, Li L, Cai Y, et al. Dynamic changes in anti-SARS-CoV-2 antibodies during SARS-CoV-2 infection and recovery from COVID-19. Nat Commun. 2020 Nov 27;11(1):6044.

22. Wu J, Liang B, Chen C, Wang H, Fang Y, Shen S, et al. SARS-CoV-2 infection induces sustained humoral immune responses in convalescent patients following symptomatic COVID-19. Nat Commun. 2021 Mar 22;12(1):1813.

23. Mahallawi W, Alzahrani M, Alahmadey Z. Durability of the humoral immune response in recovered COVID-19 patients. Saudi J Biol Sci. 2021 Feb 16.

24. Schaffner A, Risch L, Weber M, Thiel S, Jungert K, Pichler M, et al. Sustained SARS-CoV-2 nucleocapsid antibody levels in nonsevere COVID-19: a population-based study. Clin Chem Lab Med. 2020 Nov 19;59(2):e49–e51.

25. Khoury DS, Cromer D, Reynaldi A, Schlub TE, Wheatley AK, Juno JA, et al. Neutralizing antibody levels are highly predictive of immune protection from symptomatic SARS-CoV-2 infection. Nat Med. 2021 May 17.

26. Gaebler C, Wang Z, Lorenzi JCC, Muecksch F, Finkin S, Tokuyama M, et al. Evolution of antibody immunity to SARS-CoV-2. Nature. 2021 Mar;591(7851):639–44.

