## Supplemental Tables S1-S3 Figures S1-S2 for "Stable IgG-antibody levels in patients with mild SARS-CoV-2 infection"

Folkhälsomyndigheten

### Appendix

**Figure S1**. Questions used for selection of participants.

1. What County do you live in?
2. Age group and gender question.
3. Did you have symptoms of respiratory tract infection during 2020? (Yes/No).
    3.1. If Yes, specify:
        3.1.1. Mild cold, cough, rhinitis (no fever)
        3.1.2. Influenza-like symptoms including fever, muscle/joint pain, cold
        3.1.3. Sore throat with or without fever (no rhinitis or cough)
        3.1.4. Free text option
4. Did you experience symptoms during 2020 that you either suspect or have confirmed were due to COVID-19? (Yes/No)
    4.1. If Yes, specify:
        4.1.1. When did the symptoms start? (date)
        4.1.2. Have you been tested for COVID-19 with sampling in the nose or throat (PCR)? (Yes/No)
            4.1.2.1. If Yes, when was the test conducted? (date).
        4.1.3. How ill were you?
            4.1.3.1. Mild (stayed at home)
            4.1.3.2. Moderate (hospital stay)
            4.1.3.3. Severe (hospital, ICU admission)
        4.1.4. For how long period were you ill?
            4.1.4.1. Less than 10 days
            4.1.4.2. 10-20 days
            4.1.4.3. More than 20 days
        4.1.5. Have you been tested positive for antibodies against SARS-CoV-2 during 2020 (blood sampling)? (Yes/No)
            4.1.5.1. If Yes, when was the test conducted (date)?
5. Contact information and End of questions.

**Table S1.** Summary of questionnaire results used during the recruitment of study participants (see figure S1). All data are self-reported.

| | No. of presumptive positives (%)* | No of presumptive negatives (%)† | Total (%) | Presumptive pos. sampled (%) | Presumptive neg. sampled (%) | Total sampled (%) |
|---|---|---|---|---|---|---|
| **N** | 1729 (32) | 3715 (68) | 5444 (100) | 205 (42) | 279 (58) | 484 (100) |
| **Gender** | | | | | | |
| Female | 1036 (60) | 2033 (55) | 3069 (56) | 122 (60) | 153 (55) | 275 (57) |
| Male | 690 (40) | 1678 (45) | 2368 (43) | 83 (40) | 126 (45) | 209 (43) |
| Other | 3 (0.2) | 4 (0.1) | 7 (0.1) | 0 (0) | 0 (0) | 0 (0) |
| **Age group** | | | | | | |
| 18-30 | 230 (13) | 384 (10) | 614 (11) | 23 (11) | 53 (19) | 76 (16) |
| 31-50 | 876 (51) | 1767 (48) | 2643 (49) | 94 (46) | 101 (36) | 195 (40) |
| 51-70 | 592 (34) | 1460 (39) | 2052 (38) | 78 (38) | 84 (30) | 162 (34) |
| >70 | 31 (1.8) | 104 (2.8) | 135 (2.5) | 10 (4.9) | 41 (15) | 51 (11) |
| **County** | | | | | | |
| 1 | 1036 (60) | 1749 (47) | 2785 (51) | 80 (39) | 148 (53) | 228 (47) |
| 2 | 397 (23) | 1456 (39) | 1853 (34) | 71 (35) | 93 (33) | 164 (34) |
| 3 | 296 (17) | 510 (14) | 806 (15) | 54 (26) | 38 (14) | 92 (19) |
| **Symptoms of respiratory-tract infection** | | | | | | |
| Yes | 1278 (74) | 1259 (34) | 2537 (46) | 163 (80) | 88 (32) | 251 (52) |
| No[2] | 451 (26) | 2456 (66) | 2907 (53) | 42 (20) | 191 (68) | 233 (48) |
| **Symptoms of COVID-19** | | | | | | |
| Yes | 1729 (100) | 0 (0) | 1729 (32) | 205 (100) | 0 (0) | 205 (42) |
| No | 0 (0) | 3715 (100) | 3715 (68) | 0 (0) | 279 (100) | 279 (58) |
| **Symptom severity** | | | | | | |
| Mild | 1667 (96) | 0 (0) | 1667 (31) | 181 (88) | 0 (0) | 181 (37) |
| Moderate | 49 (2.8) | 0 (0) | 49 (0.9) | 19 (9.3) | 0 (0) | 19 (3.9) |
| Severe | 13 (0.8) | 0 (0) | 13 (0.2) | 5 (2.4) | 0 (0) | 5 (1.0) |
| No data | 0 (0) | 3715 (100) | 3715 (68) | 0 (0) | 279 (100) | 279 (58) |
| **Tested PCR-pos.** | | | | | | |
| Yes | 482 (28) | 0 (0) | 482 (8.9) | 162 (79) | 0 (0) | 162 (33) |
| No | 1247 (72) | 3715 (100) | 4962 (91) | 43 (21) | 279 (100) | 322 (67) |
| **Tested positive for antibodies** | | | | | | |
| Yes | 744 (43) | 0 (0) | 744 (14) | 84 (41) | 0 (0) | 84 (17) |
| No | 985 (57) | 3715 (100) | 4700 (86) | 121 (59) | 279 (100) | 400 (83) |

*Candidates that have answered “yes” on at least one of the questions on positive PCR test result, positive antibody test or symptoms of COVID-19

†Candidates that have answered “no” on at all of the questions on positive PCR test result, positive antibody test or symptoms of COVID-19

**Table S2.** Reported symptoms of respiratory tract infection among patient-cases and non-cases**.**

| **Symptoms*** | **Patient-cases (%) (n= 115)** | **Non-Cases (%) (n=127)** |
|---|---|---|
| Mild cold, cough, rhinitis (no fever) | 19 (17) | 62 (49) |
| Influenza-like symptoms including fever, muscle/joint pain, cold | 81 (70) | 49 (39) |
| Sore throat with or without fever (no rhinitis or cough) | 14 (12) | 30 (24) |
| Other, free text option | 12 (10) | 9 (7.1) |

*Symptoms were collected according to those described in Figure S1, 3.1.1-3.1.4. Participants were able to answer up to all four available options. Thus, total numbers of symptoms do not match the number of patient-cases or non-cases.

**Figure S2**. IgG levels against spike and nucleocapsid in different age groups.

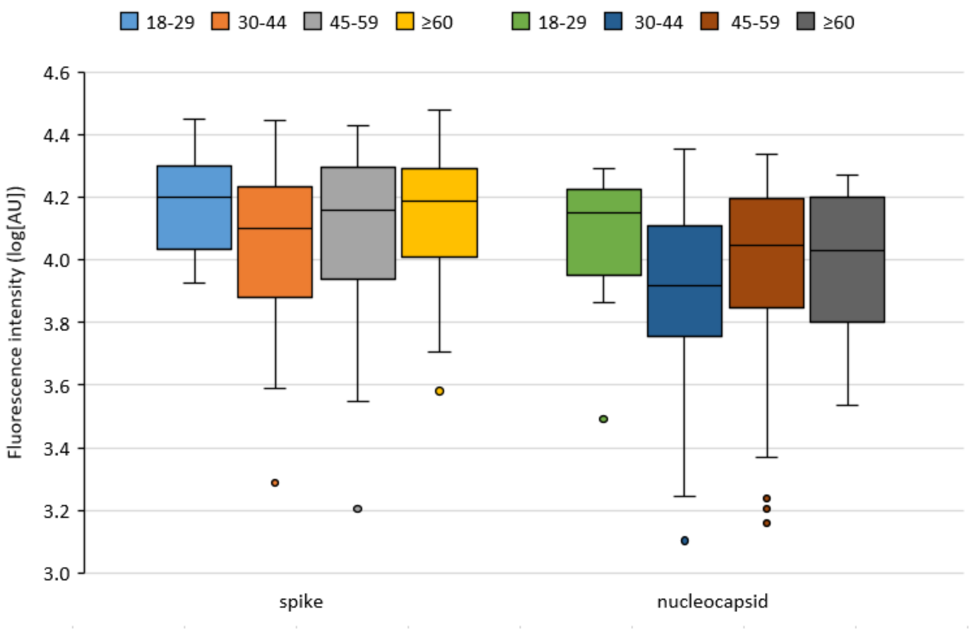

**Table S3**. Antibodies detected in serum samples of patient-cases and non-cases in the three counties.

| Reactive antibody | Cases (%) | Non-cases (%) | Non-cases, -PCR (%)* | Non-cases, RTI-symptoms (%)*,† | Non-cases, COVID-19 (%)*,† |
|---|---|---|---|---|---|
| **County 1** | **n=50** | **n=169** | **n=163** | **n=63** | **n=23** |
| Anti-spike IgG | 48 (96) | 14 (8.3) | 11 (6.7) | 4 (6.3) | 2 (8.7) |
| Anti-nucleocapsid IgG | 48 (96) | 13 (7.7) | 9 (5.5) | 3 (4.8) | 2 (8.7) |
| spike and nucleocapsid | 47 (94) | 9 (5.3) | 6 (3.7) | 3 (4.8) | 2 (8.7) |
| none | 1 (2.0) | 151 (89) | 149 (91) | 59 (94) | 21 (91) |
| **County 2** | **n=51** | **n=107** | **n=104** | **n=37** | **n=13** |
| Anti-spike IgG | 48 (94) | 4 (3.7) | 2 (1.9) | 0 (0) | 0 (0) |
| Anti-nucleocapsid IgG | 47 (92) | 2 (1.9) | 0 (0) | 0 (0) | 0 (0) |
| spike and nucleocapsid | 47 (92) | 2 (1.9) | 0 (0) | 0 (0) | 0 (0) |
| none | 3 (5.9) | 103 (96.3) | 102 (98) | 37 (100) | 13 (100) |
| **County 3** | **n=44** | **n=48** | **n=45** | **n=19** | **n=7** |
| Anti-spike IgG | 44 (100) | 3 (6.3) | 1 (2.2) | 1 (5.3) | 1 (14) |
| Anti-nucleocapsid IgG | 43 (98) | 3 (6.3) | 1 (2.2) | 1 (5.3) | 1 (14) |
| spike and nucleocapsid | 43 (98) | 3 (6.3) | 1 (2.2) | 1 (5.3) | 1 (14) |
| none | 0 (0) | 45 (94) | 44 (98) | 18 (95) | 6 (86) |

*Excluding non-cases that reported being tested positive for COVID-19 by PCR in the questionnaires.

†Includes only non-cases that reported RTI-symptoms or symptoms of COVID-19 during 2020.
